# Leveraging pediatric emergency visits as early signal for respiratory hospitalization forecasting

**DOI:** 10.64898/2026.02.25.26347074

**Authors:** Ana Guijarro Matos, Sylvain Benenati, Remi Choquet, Jean-Yves Lefrant, Mircea T. Sofonea

## Abstract

The COVID-19 pandemic exposed major vulnerabilities of hospital capacity and management worldwide, particularly in intensive care units (ICUs) and emergency rooms (ER), imposing prompt adaptation and resource reallocation. Although SARS-CoV-2 is no longer endangering healthcare systems, winter seasons continue to bring recurrent overload of critical care services, primarily due to respiratory infections. In France e.g., this pattern led to the reactivation of the national emergency response plan during the 2024-2025 seasonal influenza peak, highlighting the continuous need for improved predictive tools. However, forecasting hospitalization surges at a local scale remains a methodological challenge because the (very) low incidence numbers are subject to strong stochasticity and therefore require additional input of information and dedicated approaches. This study investigates the potential for early forecasting of respiratory infection peaks by analyzing ER visit trends. By clustering all-cause ER visits during the 2023–2025 winter seasons from the Nîmes University Hospital (France), we identified a strong temporal correlation between early pediatric hospitalizations (≤5 years old) and the following week’s adult hospitalization incidence for respiratory infections. The results suggest that tracking hospital admissions of pediatric ER visits, even without hospital care needs, can serve as a valuable early warning signal for upcoming peaks in respiratory-related hospitalizations. This predictive approach could improve hospital preparedness and resource management during seasonal influenza outbreaks.

**Author summary:** The epidemics of respiratory viruses present a significant challenge to hospitals in the temperate zone on an annual basis. Frequently, the hospital overload is mitigated by the late reactive allocation of human and material resources that are, hence, suboptimal. This study proposes a statistical framework to assist hospitals in anticipating bed requirements during seasonal influenza waves, despite high noise at the local level, by enhancing hospitalization forecasting with emergency room (ER) visit data. The prediction of the adult epidemic peak is possible through the analysis of the respiratory pediatric ER visits, which facilitates hospital management.

## Introduction

Respiratory infections rank among the most common and deadly diseases worldwide [1]. A wide range of pathogens cause these infections, each following a distinct seasonal pattern [2]. In temperate regions, most respiratory pathogens, such as influenza, coronavirus, and respiratory syncytial virus (RSV), circulate mostly during the fall and winter seasons (hereafter referred to as winter for the sake of concision), triggering major epidemic waves [3]. Although the distribution of the respiratory virus species and strains/types/variants differs over the years, their strong winter seasonality persists [4]. Additionally, Sabaté-Elabbadi et al. (2024) [5] reports a long-term rise of this winter respiratory epidemic among adults in France.

The impact of respiratory infections is not uniform across populations. While the circulating virus types differ between infants (≤5 years) and older adults (≥65 years) [4], both groups experience the most severe forms of disease [6]. This increased vulnerability is probably driven by weaker immune responses and a higher prevalence of decompensated comorbidities, such as asthma, chronic bronchitis, and chronic respiratory insufficiency [7].

During the COVID-19 pandemic, the non-pharmaceutical interventions reduced or even suppressed these seasonal respiratory epidemics [5, 8]. However, implementing such restrictive measures every winter is neither feasible nor socially acceptable. Other strategies to reduce the peak, such as vaccination campaigns, are therefore promoted through national recommendations targeting vulnerable populations. Despite these efforts, influenza vaccine coverage in France remains low (53.7% and 25.3% in populations ≥ and ≤ 65 year-old, respectively) [6] and its efficacy is also limited [9]. Consequently, even with preventive strategies in place, winter surges continue to strain healthcare systems.

French hospitals typically operate at an average occupancy rate of 80% in critical care units [10], leaving limited capacity to absorb these sudden increases in patient volume. As a result, winter respiratory epidemics often require the activation of hospital emergency plans, as observed during the winter of 2024–2025 [11, 12] or during the COVID-19 pandemic [13]. Therefore, being able to forecast this epidemic trajectory in advance is crucial for helping hospital management to avoid the activation of the hospital emergency plans. To this end, researchers now combine multiple data sources, including internet search [14] and wastewater surveillance [15], with machine learning and deep learning models [16], demonstrating strong performance at national and regional levels. However, forecasting at the level of a single hospital remains challenging, as stochastic noise dominates where patient flow is too small for convergence properties (e.g. law of large numbers, central limit theorem) to hold, thus hindering the use of classical forecasting methods (e.g. SARIMAX, ETS [17]).

In this context, well-documented emergency room (ER) data represent a rich, real-time source of information that can improve forecasting performance of respiratory epidemics to help guide hospital management decisions. This study, therefore, focuses on identifying patient groups presenting to the ER one week before the peak of adult respiratory infections. Anticipating this peak one week in advance would provide sufficient time for hospitals to prepare and reorganize resources to manage the incoming patient surge. The analysis was conducted at a single center (Nîmes University Hospital, in southern mainland France) and counters the aforementioned high noise-to-signal ratios with. Indeed, the hospital hosts approximately 2000 beds and 330 daily ER visits. Thus, a daily-level analysis is imperative to provide actionable insights that can support the emergency department directly.

## Materials and methods

### Data

The present study retrospectively analyzes the data prospectively collected from patients admitted to the ER of Nîmes University Hospital (Fig 1). The data is anonymous for the present study. According to the French law, the patient’s informed consent was waived, and the study was approved by the Institutional Review Board [18] (IRB: #26.02.01, Président: Pr. T. Lavabre Bertrand). The data corresponds to 243,997 emergency room daily visits between June 20th, 2023, and June 20th, 2025, at the University Hospital of Nîmes. We remove 47,265 patients whose age, sex, or primary diagnosis were not known, hence leading to an analytical sample of 196,732 patient visits. The data contains two winter peaks for adult respiratory hospitalizations: the 2024 peak from November 19, 2023, to January 26, 2024, and the 2025 peak from November 18, 2024, to January 25, 2025. These peaks correspond to the dates from three weeks before the start until the peak of the epidemics considered in Bernard-Stoecklin et al. (2025) [6]. Given the large number of different and highly specific primary and secondary diagnoses, we group them into 17 categories (15 respiratory-related S1 List, trauma and others). We also categorized the vital signs (temperature, blood pressure, pain score, oxygen saturation, and heart rate) with sub-stratification by age and sex, as shown in S1 Table. Lastly, we remove the secondary diagnosis due to its high number of missing values.

**Fig 1.**
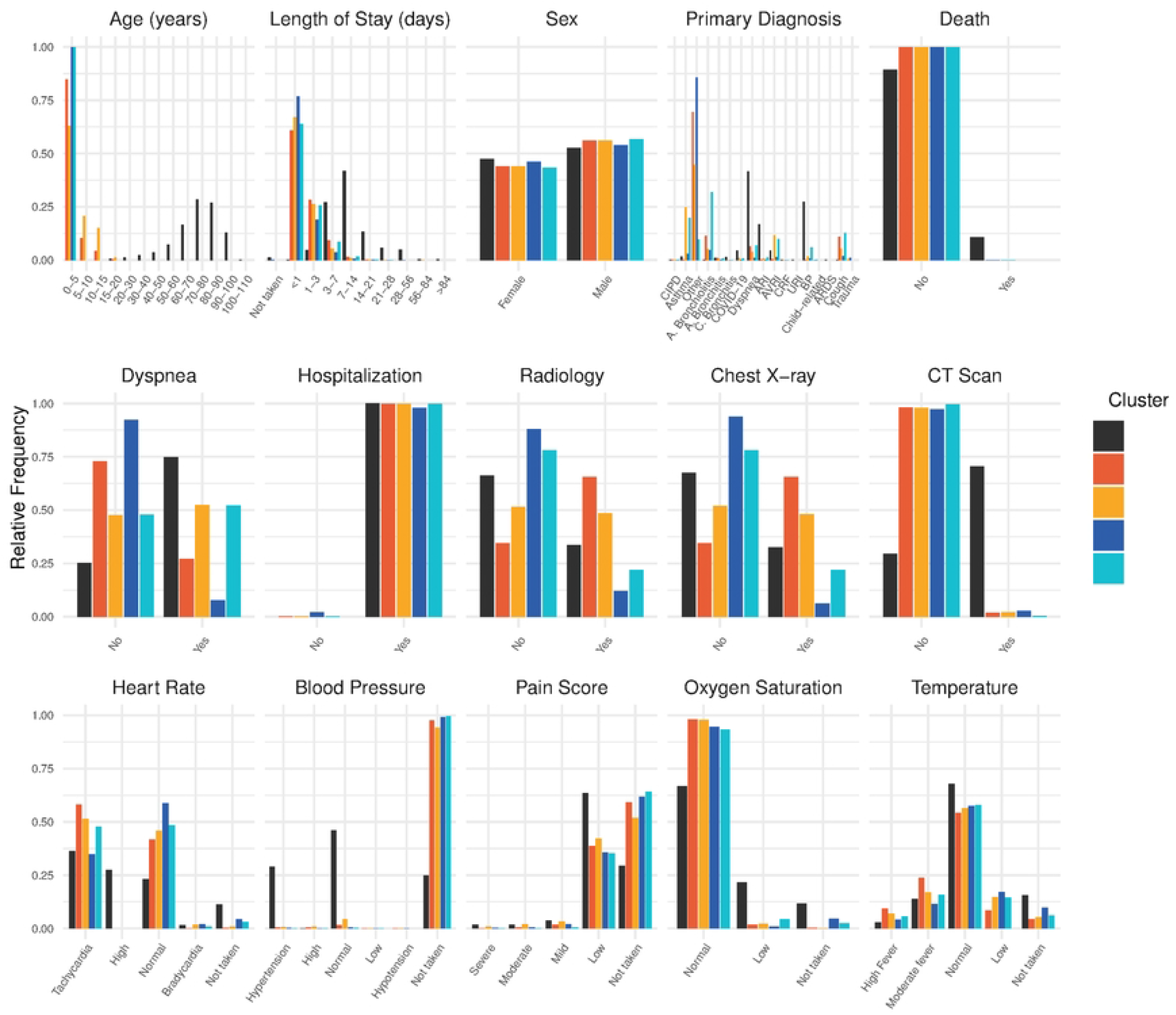
Flowchart of the methodological framework.

### Clustering

To reduce computational complexity, we perform a clustering analysis only on the winter peaks to ensure a sufficient signal-to-noise ratio. To prepare the data for clustering, we applied a Multiple Correspondence Analysis (MCA) [19] that reduces the 34 categorical variables into 4 numerical dimensions. We then apply a hierarchical consensus clustering [20] from the `sharp` package to the MCA results, which clustered the patients into 12 clusters for the 2024 peak and 15 clusters for the 2025 peak (S1 Fig). After assigning each individual to a cluster during the winter peak, we evaluate the relationship between each 7-day lagged cluster and adult respiratory infections using the Granger causality test on its 7-day moving right-averaged daily time series [21]. To extend the analysis to the entire study period, we assign a cluster to the remaining patients (outside the peak period) using a Support Vector Machine (SVM) [22] classifier with a radial kernel, trained on the winter peak clustering results for each of the two peak. We employ this supervised approach due to its capacity to learn non-linear decision boundaries in the multidimensional feature space and ensure consistent, reproducible cluster attribution across the entire dataset. Afterward, to confirm the causality of the time series over the entire period, we perform the 7-day lagged Granger test on all peaks, comparing the 7-day moving right-averaged time series of adult respiratory hospitalizations and cluster. We identify the peaks when their peakness score (computed by comparing each point value to the mean of its 14-days neighborhood, see the score type 3 in the `scorepeak` package [23] for more details) exceeds a predefined threshold.

### Daily forecast

Diverse methodological methods can be employed to forecast time series. Although classical approaches, such as ARIMAX, have been shown to perform satisfactorily, recent studies have demonstrated that deep learning techniques, particularly Long Short-Term Memory (LSTM) models, exhibit superior performance in the context of time series forecasting [24–26]. In this study, we employ a model composed of bidirectional Long Short-Term Memory (bi-LSTM) layers [26, 27], trained using the mean squared error (MSE) as the loss function. To assess the forecast uncertainty, we compute the confidence interval by generating 100 iterations with the dropout layer activated, following the Monte Carlo dropout approach [28]. To provide real-time forecasting that can support hospital management, we develop an updatable model. This model combines incremental learning through daily gradient updates on newly available observations with periodic full retraining to prevent catastrophic forgetting and maintain long-term predictive accuracy [29]. We train the model on data containing at least one peak (2024 epidemic), commencing on 18th November 2024, and test over the next 7-day. As the model is updated on a daily basis, the training set increases by one and the test dataset shifts. We emphasize the model’s performance in the test sets that included in the 2025 peak period.

In addition to comparing models trained on each cluster time series, using a seven-day input window to forecast the following seven days, we include a second feature—the seven-day lagged respiratory adult hospitalization time series—to assess whether including the target variable could enhance predictive performance. Moreover, in order to demonstrate the significance of the present study, the Naive Model (𝒩) is also considered. This model assumes that the same number of patients will arrive over the following seven days.

### Metrics

This study focuses on predicting the epidemic peak one week in advance to help anticipate hospital bed needs and optimize resource allocation at the critical period. For this reason, we calculate the metrics over the predicted horizon of each time step for every model, with particular focus on the 2025 peak period, thereby obtaining a distribution of each metric for every updatable model. To assess the accuracy of the forecast shape and magnitude, we compute the Root Mean Square Error (RMSE) and the Mean Absolute Error (MAE) [30] using the `score` Python package [31]. To evaluate the quality of the confidence intervals, we implement the Weighted Interval Score (WIS), as described in [32]. In addition, following Massey et al. (2024) [17], we utilize the Brier score, which is suitable to detect crises while forecasting due to its binary nature. We defined crisis period as time interval the cumulative sum of the three consecutive days of adult respiratory patients exceeds the 70th percentile of all three-day cumulative sums from the complete dataset, following the ER head expertise.

### Code

The following link contains the codes: https://zenodo.org/records/18712502

## Results

### Clustering

We perform clustering on each peak present in the dataset, corresponding to the 2024 and 2025 seasonal peaks of respiratory infections. In each case, the time series of one cluster is cross-correlated with adult respiratory hospitalizations (𝒜^R^) according to the Granger causality test (S2 Table). Throughout all the study period, the 2024 peak cluster has 3,912 patients (𝒫_2024_), while the 2025 peak cluster contains 1,622 patients (𝒫_2025_). Notably, both in 2024 and 2025 clusters exhibit the peaks activity one week prior to the adult respiratory hospitalization peaks. The patient-characterization profiles are highly similar across both clusters (Fig 2 and S3 Table). Patients in both clusters were predominantly pediatric (< 5 years). Infants are at high risk for respiratory infections and often act as index cases within households [33]. Hence, it is also appropriate to compare these clusters with those derived from ER visits aged 0–5 years (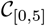, comprising 27,021 patients) and including those specifically diagnosed with respiratory conditions or dyspnea (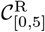, with 3,966 patients) (S3 Table).

**Fig 2.**
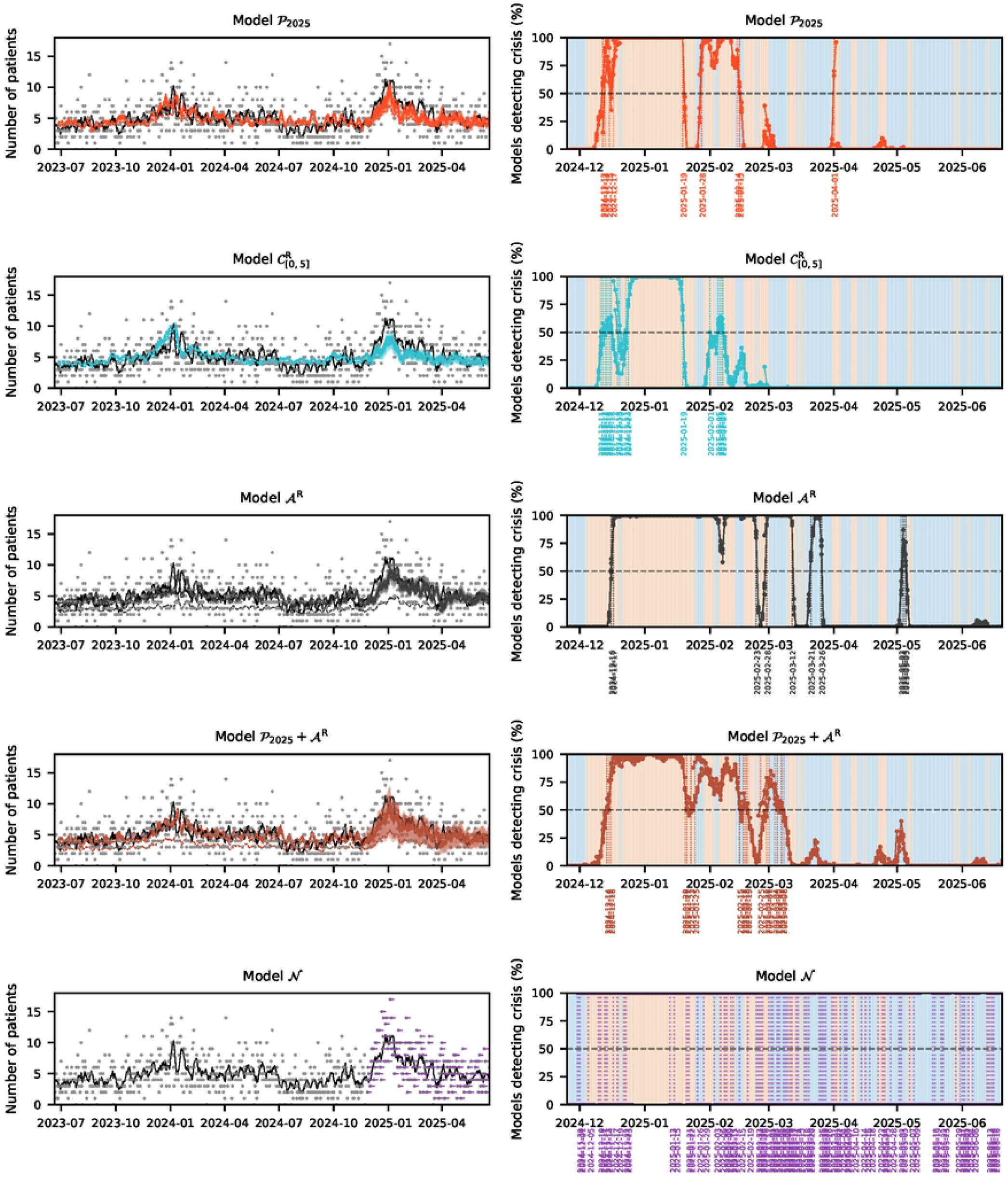
Key characteristics of the identified clusters. The bar plots provide a visual representation of the distribution of key characteristics for each cluster. The leftmost bar corresponds to 𝒜^R^, followed by the clusters obtained through the implementation of clustering and causality (𝒫_2025_, 𝒫_2024_) and finally, the simplified version 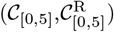. Primary diagnosis abbreviations correspond to the following: Acute Bronchiolitis (A. Bronchiolitis), Chronic Interstitial Pulmonary Disease (CIPD), Chronic Bronchitis (C. Bronchitis), Acute Bronchitis (A. Bronchitis), Acute Viral Respiratory Infection (AVRI), Chronic Respiratory Failure (CRF), Unspecified Respiratory Infection (URI), Acute Respiratory Infection (ARI), Acute Respiratory Distress Syndrome (ARDS), Bacterial Pneumonia (BP).

These four clusters (𝒫_2025_, 𝒫_2024_, 𝒞_[0,5]_, and 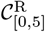) exhibited very similar characteristics, as shown in Fig 2. There were no sex-based differences, and the hospital stays were short. At least 89% of patients in each cluster were hospitalized for less than three days. Despite these overall similarities, the clusters differ in specific variables, including diagnosis, radiography, and symptoms of dyspnea. Indeed, except for the 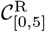 cluster, characterized by asthma (20%), acute bronchiolitis (32%), and cough (13%), the predominant diagnoses in the other clusters were non–respiratory-related (“other” diagnosis for 45% of patients in the 𝒫_2024_ cluster, 69% in the 𝒫_2025_ cluster, and 86% in the *C*_[0,5]_ cluster). Radiography use is substantially higher in the peak clusters (48% in *P*_2024_ and 66% in 𝒫_2025_) compared with the other two, where only 12% of patients in the 𝒞_[0,5]_ cluster and 22% in the 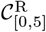 cluster underwent imaging. Dyspnea symptoms are predominant in patients from the 𝒫_2024_ cluster (52%) and in patients from the 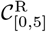 cluster (52%), but are less frequent in the 𝒫_2025_ cluster (27%) and in the 𝒞_[0,5]_ cluster (7.7%).

### Daily forecast

We compare the 10 different models, the naive model (𝒩), the five models trained on the time series of the four clusters obtained (𝒫_2024_, 𝒫_2025_, 𝒞_[0,5]_, and 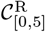), the model trained with the 𝒜^R^ time series itself and the four models that incorporating the 𝒜^R^ time series into the four cluster-based models (𝒫_2024_ +𝒜^R^, 𝒫_2025_ +𝒜^R^, 𝒞_[0,5]_ +𝒜^R^, and 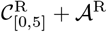) (Fig 3, S2 Fig). Across all bi-LSTM models, the forecasts of respiratory infection epidemics reveals a high degree of similarity. This forecasting model accurately predicts a respiratory peak between December and February 2025. In all this models, the predicted amplitudes are lower than the observed values, even if the forecast is closer to the 7-day moving right-average. In contrast, the model 𝒩 did not generate the peak shape, it replicated only the results obtained on the day.

**Fig 3.**
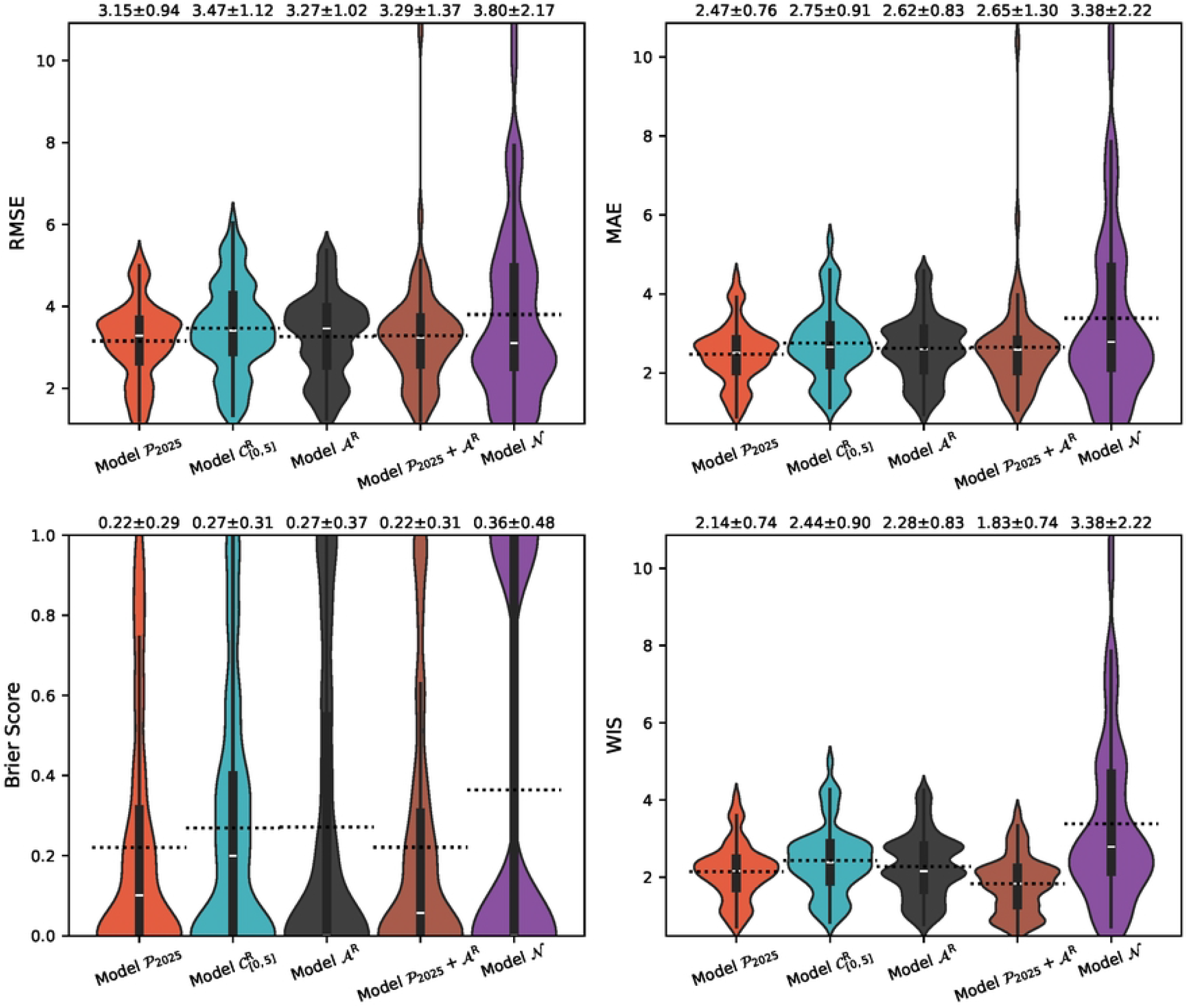
Daily forecast of adult respiratory hospitalizations. In the left panel, the forecasts of model 𝒩 and models generated using the updatable bi-LSTM model trained on different time series: 𝒫_2025_, 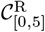, 𝒜^R^ and 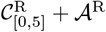. All input series are incorporated with a 7-day lag to predict one week ahead. The solid colored line represents the median forecast, with shaded areas indicating the corresponding confidence interval. The dashed lines correspond to the prediction of the training sets. The black line shows the 7-day moving right-average of observed adult respiratory hospitalizations, while gray dots denote the daily observed data. The right panel represents the proportion of iterations of each model (𝒫_2025_, 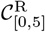, 𝒜^R^ and 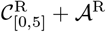, 𝒩) that detect a crisis per day, during the period in which the models were tested. The vertical lines correspond to the day that the model will declare a crisis or remove the crisis, since there is at least 50% of iterations detecting a crisis that day, with a dashed line corresponding to this threshold of 50% iterations. The graph is colored with the real crisis detection, where the orange zone correspond to the crisis days and the blue area the non crisis days.

The model 𝒜^R^ produces the most comparable peak shape; however, it exhibits a one-week delay. This temporal shift occurs similarly when the time series is incorporated into the training process (Fig 3, S4 Fig). In fact, crisis detection for models incorporating 𝒜^R^ is delayed by at least two days compared to model versions without 𝒜^R^, as shown in Fig 3. Models without 𝒜^R^ achieve earlier crisis detection and reduced false positive rates during non-crisis periods (S5 Table). Conversely, the model 𝒩 exhibits variable crisis detection performance depending on the daily observed data.

Quantitatively, performance metrics, provided in S4 Table, all show a slight mean variation across all the predicted period for the simple models RMSE: 0.15, MAE: 0.16, Brier score: 0.09, WIS: 0.14, and this difference is reduced when we add the 𝒜^R^ (RMSE: 0.04, MAE: 0.07, Brier score: 0.02, WIS: 0.06). Overall, the model 𝒫_2025_ achived the lowest mean of the metrics sum, with a mean difference of 2.83 from the highest, the model 𝒩. However, when restricting the comparison to trained models only, the differences were substantially smaller. The mean difference of the metrics sum is 0.48 against the 𝒞_[0,5]_, and 0.44 against the model 𝒜^R^.

The period used to test the models is quite large, while for the hospital, only the peak period and the determination of the crisis period are of importance. For that purpose, the same metrics are calculated over the peak period and outside this period (S3 Fig). The distribution of metrics outside the crisis period does not demonstrate variation between the models. In contrast, during the peak period a slight variation is shown (Fig 4). During this period, the model 𝒞_[0,5]_ obtains the higher mean value for all metrics (RMSE: 3.56, MAE: 2.84, Brier score: 0.35, WIS: 2.50). Conversely, the model𝒫_2025_ achieves the lowest mean value for all metrics (RMSE: 3.15, MAE: 2.47, Brier score: 0.22, WIS: 2.14). The incorporation of 𝒜^R^ into the aforementioned models led to a deterioration in the RMSE and MAE, particularly the standard deviation. Specifically, RMSE increased from 3.15 ± 0.094 and MAE from 2.47 ± 0.76 for the model [0,5] to 3.29 ± 1.37 and 2.65 ± 1.30, respectively, for the model 𝒫_2025_ + 𝒜^R^, as illustrated in Fig. 4. This effect is analogous in the rest of the model. Although the inclusion of 𝒜^R^ slightly reduces the mean sum of metrics over the entire test period (see S4 Table), this apparent improvement is accompanied by increased dispersion during the peak period. Indeed, the distributions of RMSE and MAE of these models exhibit a pronounced violin shape, with a substantial presence of extreme high values. This high variability is likewise observed for the baseline model 𝒩.

**Fig 4.**
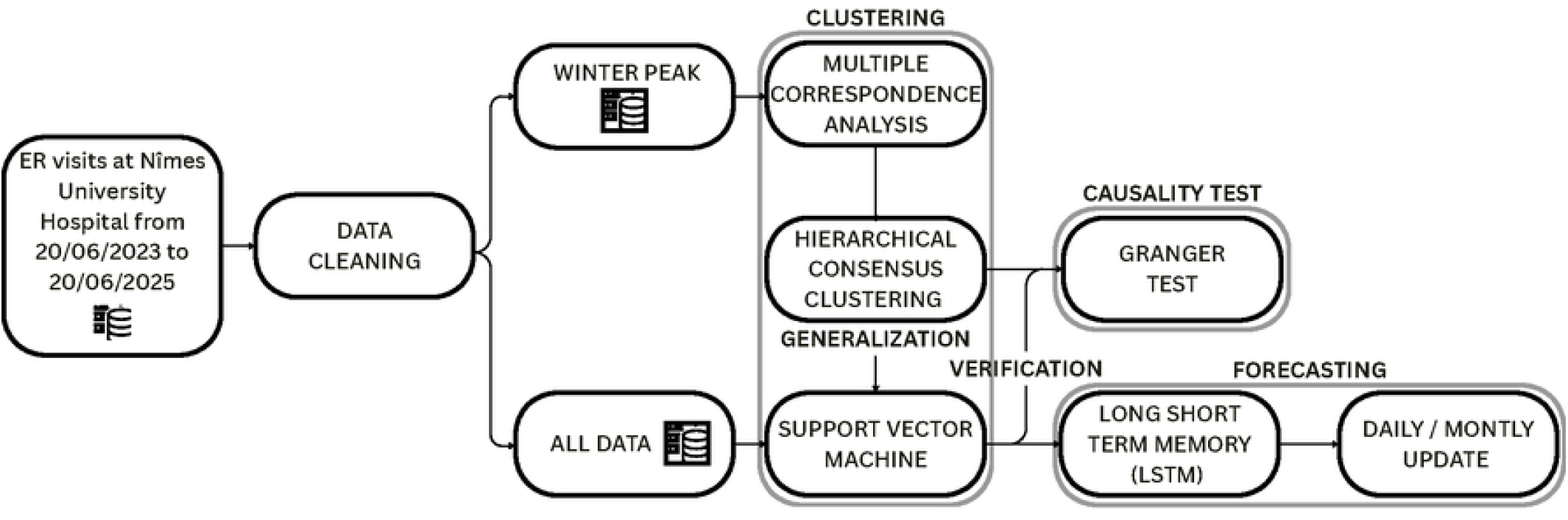
Model metrics for adult respiratory hospitalizations forecasts over the 2025 peak period. Violin plots showing the distribution of four performance metrics (RMSE, MAE, Brier score, WIS) obtained with the updatable bi-LSTM model trained on the time series 𝒫_2025_, 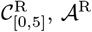 and 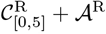 and the naive model *N*. The mean ± standard deviation is shown at the top of each panel, with the dashed line indicating the mean.

## Discussion

The clustering of ER visits during the two seasonal respiratory epidemics (2023-2024, 2024-2025) yielded consistent results, with comparable patient characteristics. In both years, the cluster cross-correlated with respiratory adult hospitalization is predominantly composed of pediatric patients. Despite minor variations in the distribution of specific characteristics, including diagnoses categories, imaging, and dyspnea indicators, the majority of features exhibited comparable distributions across the two seasons. This temporal consistency suggests that the identified pediatric cluster captures a stable epidemiological signal rather than season-specific noise. The observation that pediatric ER visits exhibit a strong cross-correlation with adult respiratory hospitalizations is not unexpected. This phenomenon can be explained by the well-established fact that children are at a higher risk of respiratory infections and frequently act as index case of respiratory viruses within households [33]. It is essential to note that our findings support the notion that pediatric ER visitation patterns can serve as effective one-week warning signals for the subsequent surge in adult hospital burden.

Extending this observation, we perform a characterization of the clusters of all children aged 0–5 years visiting the ER and those with respiratory-related diagnosis within the same age range. The distributions of the majority of the characteristics exhibit a high degree of similarity between these two clusters and the previously identified ones. This finding suggests that a simplified clustering approach, focusing exclusively on respiratory ER visits among pediatric patients, may sufficiently capture the relevant epidemiological dynamics. This simplification is particularly relevant for real-world implementation, as it reduces computational complexity and data requirements, thereby facilitating integration into hospital surveillance workflows.

The forecasts demonstrate notable similarities across all models, with all models exhibiting superior performance in comparison to the baseline model 𝒩, which remains constant each day. Consequently, the decision-making using model 𝒩 is ineffective and inadequate. This underscores the importance of the study and serves to affirm that any of the models will constitute a substantial improvement. The models trained on the four clusters (𝒫_2024_, 𝒫_2025_, 𝒞_[0,5]_, and 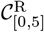) perform satisfactorily in forecasting winter epidemic peaks. Incorporating 𝒜^R^ time series as a predictor resulted in a consistent degradation of model performance. In particular, the model 𝒜^R^, which utilizes lag-7 adult hospitalizations to predict future values, demonstrated the poorest performance. Indded, a visual inspection of the forecast reveals a systematic delay of approximately seven days, suggesting that the model has learned a lagged replication of the same signal rather than a true predictive relationship. As demonstrated in Fig 3, the delay patterns exhibited by models incorporating 𝒜^R^ as an input are analogous. This phenomenon is substantiated by the crisis detection (Fig 3, S4 Fig), where these models, including 𝒜^R^ time series, start detecting the crisis days after the rest of the models.

While this late detection results in greater temporal stability and consequently yields lower mean Brier scores, the critical consideration for hospital operations is the ability to prepare adequately for impending crises. Hence, these models may offer retrospective validation rather than prospective warnings, thereby limiting their practical applications in decision-making processes related to resource allocation. The models demonstrate remarkable consistency with similar performance metrics, obtaining superior performance with the model 𝒫_2025_. However, the differences are negligible in practical terms. The alignment of results across the diverse models indicates that the forecasting framework exhibits considerable robustness and is not dependent on the specific cluster. However, when considering the operational implementation of routine hospital surveillance systems, the respiratory-diagnosed 0–5-year-old age group emerges as the optimal choice to train the model. The validity of this recommendation is supported by two key factors – first, the methodological parsimony and ease of implementation of the model 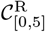. Secondly, the model 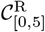 exhibited superior sensitivity in detecting crisis events, exhibiting a lower mean Brier score and reduced variability when compared to the model 𝒞_[0,5]_. The integration of practical simplicity and improved crisis detection capability suggests that clustering pediatric ER visits with respiratory diagnosis is particularly well-suited for incorporation into epidemiological surveillance programs.

The present study has its limitations. Firstly, the study was conducted in a single center, where the relatively low number of patients results in a high signal-to-noise ratio. Consequently, forecasting errors frequently correspond to small absolute patient counts, which may disproportionately affect performance metrics. However, this limitation also underscores the practical relevance of the study, as hospital-level decision-making frequently relies on this data and is more effective than global scale [34]. Secondly, the deep learning models were trained on a restricted dataset comprising only two epidemic seasons, one for training (2024 peak) and one for testing (2025 peak). This restricted temporal scope imposes limitations on the models’ capacity to discern a broader range of epidemic patterns, a limitation that may compromise the generalizability of the models’ predictions. This is a recognized challenge faced by recurrent neural networks when trained on short time series data. To increase the precision and robustness of the model, it is necessary to incorporate additional seasons of data. Thirdly, it is important to consider that model parametrization is inherently context-specific. Deep learning models require recalibration for each hospital setting, as variations in patient populations, organizational structures, and care pathways limit direct transferability. As such, the present study should be understood as a proof of concept.

Overall, the primary objective of this research is to provide direct assistance to hospitals in the development and implementation of data-driven early warning systems. The demonstration of the predictive value of pediatric respiratory ER visits provided support for the integration of this model into management routines in hospitals. The objective of this integration is to enhance patient care, optimize resource allocation, and elevate preparedness during seasonal respiratory epidemics. Consequently, subsequent studies will prioritize the validation of these findings during both prior and upcoming epidemic seasons, as well as the development of an operational platform for real-time implementation within hospital systems. Ultimately, re-parameterizing the models is essential for facilitating their implementation in other hospitals with analogous organizational structures.

## Conclusion

Seasonal respiratory infections have been identified as the primary cause of the observed excess in hospitalizations during the winter months in France [6]. This phenomenon has seen a marked intensification recently, as evidenced by the increasing magnitude of hospitalization peaks over time [5]. Despite the general awareness among healthcare professionals regarding this persistent challenge, the variability in circulating viruses and the annual fluctuations in epidemic timing complicate the anticipation and allocation of resources [2]. In this context, the present study demonstrates that the ER pediatric visit time series is the only endogenous indicator that consistently exhibits an informative cross-correlation at a 7-day lag with adult respiratory hospitalizations.

Specifically, visits from children aged 0–5 years with respiratory diagnosis have been identified as a leading early signal. Consequently, we developed a bi-LSTM prediction model trained with the time series of this cluster. This model demonstrates the capacity to predict adult respiratory hospitalizations with one week of lead time at a single hospital center. The integration of this predictive model into hospital operations has the potential to enhance operational preparedness and resource allocation.

## Data Availability

Individual-level data cannot be made publicly available in order to ensure compliance with data protection regulations and to safeguard participant confidentiality. The codes are publicly available on Zenodo (link provided in the Main Text).

https://zenodo.org/records/18712502

## Supporting information

**S1 Table. Vital sign classification as a function of age and sex**. Categorization of the vital signs (oxygen saturation, temperature, blood pressure, pain score, heart rate) of each of the patients depending on their age and sex.

**S1 List. Respiratory diagnoses classification**. The substantial number of different diagnoses complicates the process of clustering. To address this challenge, we relied on professional advice to categorize all the diagnoses with any respiratory characteristic into 15 categories (Asthma, Child-related, Acute Bronchiolitis (A. Bronchiolitis), Chronic Interstitial Pulmonary Disease (CIPD), Chronic Bronchitis (C. Bronchitis), Acute Bronchitis (A. Bronchitis), Acute Respiratory Distress due to Symptomatic COVID (COVID-19), Acute Viral Respiratory Infection (AVRI), Chronic Respiratory Failure (CRF), Unspecified Respiratory Infection (URI), Cough, Dyspnea, Acute Respiratory Infection (ARI), Acute Respiratory Distress Syndrome (ARDS), Bacterial Pneumonia (BP)).

**S1 Fig. Metrics of hierarchical clustering as a function of the number of clusters**. The left-hand plot corresponds to the result for clustering the 2024 peak, while the right-hand plot corresponds to the cluster of the 2025 peak. To determine the optimal number of clusters, several metrics are evaluated, with the aim of minimizing both relative and absolute loss. This approach reflects a balance between cluster compactness and interpretability. Furthermore, the number of clusters should be sufficiently large to ensure that each cluster contains sufficient information.

**S2 Table. Patient distribution across clusters with Granger test p-values**. The table presents the number of patients per cluster after clustering over the two peak periods, along with their respective p-values associated with the lag-7 cross-correlation Granger test with adult respiratory hospitalizations. Cluster 1 corresponds to the entire cohort of ER visits, devoid of any specific characterization; the remaining clusters are smaller and more specific.

**S3 Table. Distribution of patient characteristics per cluster**. For each cluster, the table reports the absolute number of patients exhibiting each characteristic, along with the corresponding percentage within that cluster.

**S2 Fig. Model forecast**. Prediction generated using the baseline model (𝒩) and the updatable bi-LSTM model trained on multiple time series: 𝒫_2024_, 𝒫_2025_, 𝒞_[0,5]_, 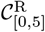, 𝒜^R^, 𝒫_2024_ +𝒜^R^, 𝒫_2025_ +𝒜^R^, 𝒞_[0,5]_ +𝒜^R^, and 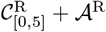. All input series are incorporated with a 7-day lag to predict one week ahead. The solid-colored line in the graph represents the median forecast, with the shaded areas indicating the corresponding confidence interval. The dashed lines represent the prediction of the training sets. The black line shows the 7-day moving right-average of observed adult respiratory hospitalizations, while gray dots denote the daily observed data.

**S4 Table. Metrics table over all the test period**. The table presents the mean and standard deviation of the metrics (RMSE, MAE, Brier score, and WIS), as well as the sum of the metrics for all models across all test periods.

**S3 Fig. Violin plot of the metrics in function of the model and whether is peak period or not**. Representation of the metrics distribution per model, considering the test set’s timing relative to the peak period. The mean ± standard deviation is shown at the top of each violin, with the dashed line indicating the mean.

**S4 Fig. Percentage crisis detection per model**. Representation of the proportion of iterations of each model that detected a crisis per day during the period in which the models were tested. The vertical lines correspond to the day that the model will declare a crisis or remove the crisis, since there is at least 50% of iterations that detect a crisis on that day. The figure displays a dashed line corresponding to this threshold of 50% iterations. The graph is colored according to the real crisis detection, where the orange zone correspond to the crisis days and the blue area the non crisis days.

**S5 Table. Confusion matrix for each model stratified by peak period**. Mean ± standard deviation of the percentage of true negatives (TN), false positives (FP), false negatives (FN), and true positives (TP) for all model predictions, separately evaluated during peak and non-peak periods.

## Acknowledgments

AGM, JYL, RC and MTS acknowledge funding from the ExposUM Institute of the University of Montpellier (funded by the Agence Nationale de la Recherche, ANR-21-EXES-0005, part of the France 2030 programme, and by the Occitanie Region ; NEXUS EMIPSA project). MTS also acknowledge funding from the ANRS Maladies Infectieuses Émergentes and France 2030/SGPI (ANRS-24-PEPRMIE-0003 PreViX project).

